# Genetic associations among internalizing and externalizing traits with polysubstance use among young adults

**DOI:** 10.1101/2023.04.04.23287779

**Authors:** Leslie A Brick, Chelsie E Benca-Bachman, Emma C Johnson, Daniel E. Gustavson, Matthew Carper, Rohan HC Palmer

## Abstract

Though most genetic studies of substance use focus on specific substances in isolation or generalized vulnerability across multiple substances, few studies to date focus on the concurrent use of two or more substances within a specified time frame (i.e., polysubstance use; PSU). We evaluated whether distinct genetic factors underlying internalizing and externalizing traits were associated with past 30-day PSU above variance shared across general psychopathology and substance use (SU). Using Genomic Structural Equation Modeling, we constructed theory-driven, multivariate genetic factors of 16 internalizing, externalizing, and SU traits using genome-wide association studies (GWAS) summary statistics. Next, we fit a model with a higher order SU-related psychopathology factor as well as genetic variance specific to externalizing and internalizing (i.e., residual genetic variance not explained by SU or general psychopathology). GWAS-by-subtraction was used to obtain single nucleotide polymorphism effects on each of these factors. Polygenic scores (PGS) were then created in an independent target sample with data on PSU, the National Longitudinal Study of Adolescent to Adult Health. To evaluate the effect of genetic variance due to internalizing and externalizing traits independent of variance related to SU, we regressed PSU on the PGSs, controlling for sex, age, and genetic principal components. PGSs for SU-related psychopathology and non-SU externalizing traits were associated with higher PSU factor scores, while the non-SU internalizing PGS was not significantly associated with PSU. In total, the three PGSs accounted for an additional 4% of the variance in PSU above and beyond a null model with only age, sex, and genetic principal components as predictors. These findings suggest that there may be unique genetic variance in externalizing traits contributing to liability for PSU that is independent of the genetic variance shared with SU.

## Introduction

A growing body of evidence highlights the adverse effects of multiple and co-occurring substance use disorders (SUDs) on individuals, including increased risk for emergency hospitalization (1), greater disease burden (2), and more severe comorbidities with greater service use (3). Comorbidity among SUDs is common: 15% of individuals diagnosed with past year alcohol use disorder were also diagnosed with at least one other SUD while 56.8% of those with opioid use disorder were also diagnosed with at least one other SUD (4). While much of the prior research in substance use focuses on misuse of single substances, often in isolation, many users do report use of more than one substance in a given period of time. In fact, the problematic use of multiple substances may exacerbate or contribute to increased severity of other psychiatric concerns, with individuals who exhibit multiple SUDs presenting with higher prevalence of mood, personality, and posttraumatic stress disorders than those diagnosed with a single SUD (4). Furthermore, the combination, or mixing, of multiple substances can result in serious health problems, as nearly half of all drug overdose deaths reported in the United States in 2019 involved multiple drugs (5).

Polysubstance use (PSU) is defined broadly as the use of two or more substances within a defined period, either simultaneous or on separate occasions (6, 7). PSU may be intentional or unintentional and has been associated with greater risk for SUD (8), as well as lifetime suicide attempts (3), and having experienced childhood maltreatment (9). Early adolescent (i.e., prior to age 16) polysubstance use of cannabis, cigarettes, and alcohol was associated with substance use problems and SUD in early adulthood (10). Furthermore, polysubstance using youth were found to be more likely to report psychological distress (11), depressive symptoms (12), delinquency (13), and risky sexual behaviors (14).

Indeed, a multitude of pathways may potentiate the risk for problematic substance use and risk for development of SUD, with two central domains including externalizing (i.e., behavioral disinhibition, poor self-regulation, aggression) and internalizing (i.e., negative affect, anxiety) problems. Both internalizing and externalizing traits play significant roles in the development of adolescent substance use (15, 16, 17). Authors of a recent study using network analysis to examine comorbidity between substance use behaviors and mental health found evidence for a separate clustering of substance use behaviors, internalizing symptoms, and externalizing symptoms as well as connections between individual substances and internalizing/externalizing (18). Specifically, they found that substance use behaviors were strongly associated with each other and that the use of cannabis, alcohol, and prescription drugs were associated with externalizing symptoms while prescription drugs were also associated with internalizing symptoms. Together, the extant research seemingly supports that both internalizing and externalizing play an important role in substance use, yet few studies have distinguished PSU as a distinct substance use-related phenotype.

While substance use disorders are etiologically complex -- and influenced by both genetic and environmental (non-genetic) factors -- twin and molecular genetic studies have consistently supported a moderate heritability for a shared vulnerability across multiple substances (i.e., common liability)(19, 20). Palmer et al. (2015) estimated that 20% of the variance in general liability to substance dependence (e.g., cannabis, alcohol, nicotine, cocaine, or other illicit drugs) was attributed to common single nucleotide polymorphisms (SNPs), evidenced through the use of genome-based restricted maximum likelihood estimation of SNP-level data from the Study of Addiction: Genetics and Environment. Similarly, a recent investigation using genomic structure equation modeling (SEM) to leverage genome-wide association study (GWAS) summary statistics across numerous large studies revealed that common genetic liability across four substance use phenotypes (problematic alcohol use, problematic tobacco use, cannabis use disorder, and opioid use disorder) was associated with several behavioral traits consistent with stage-based facets of addiction, such as risk taking, executive function, and neuroticism (21). Indeed, these findings are consistent with evidence that genetic correlations among substance use phenotypes, as well as between substance use phenotype and several psychiatric traits, reveal a pattern of common risk or vulnerability (22). Still, few studies have examined genetic variance related to PSU, instead focusing on *lifetime* vulnerability to multiple SUD.

While no large-scale studies exist to directly examine the genetic architecture of PSU using GWAS, we utilized publicly available data to investigate whether genetic variance in substance use behavior and internalizing/externalizing traits explain variation in liability for PSU behavior. To accomplish this, we used summary statistics from large-scale genome-wide association studies (GWAS) to construct theory-driven, multivariate genetic factors encompassing the most salient internalizing and externalizing traits relevant to risk for substance use. We then identified genetic variance in internalizing and externalizing that is independent of substance use behaviors using genome wide association-by-subtraction (23), and created polygenic scores representing internalizing traits not related to substance use, externalizing traits not related to substance use, and substance use-related traits (including variance shared across substance use, internalizing, and externalizing psychopathology). Finally, we evaluated whether these polygenic scores conferred risk for polysubstance use in young adults using a large, independent, nationally representative sample of young adults in the United States.

## Methods

Please see **Figure 1** for an overview of methods and Supplementary Materials for additional information throughout.

**Figure 1.**
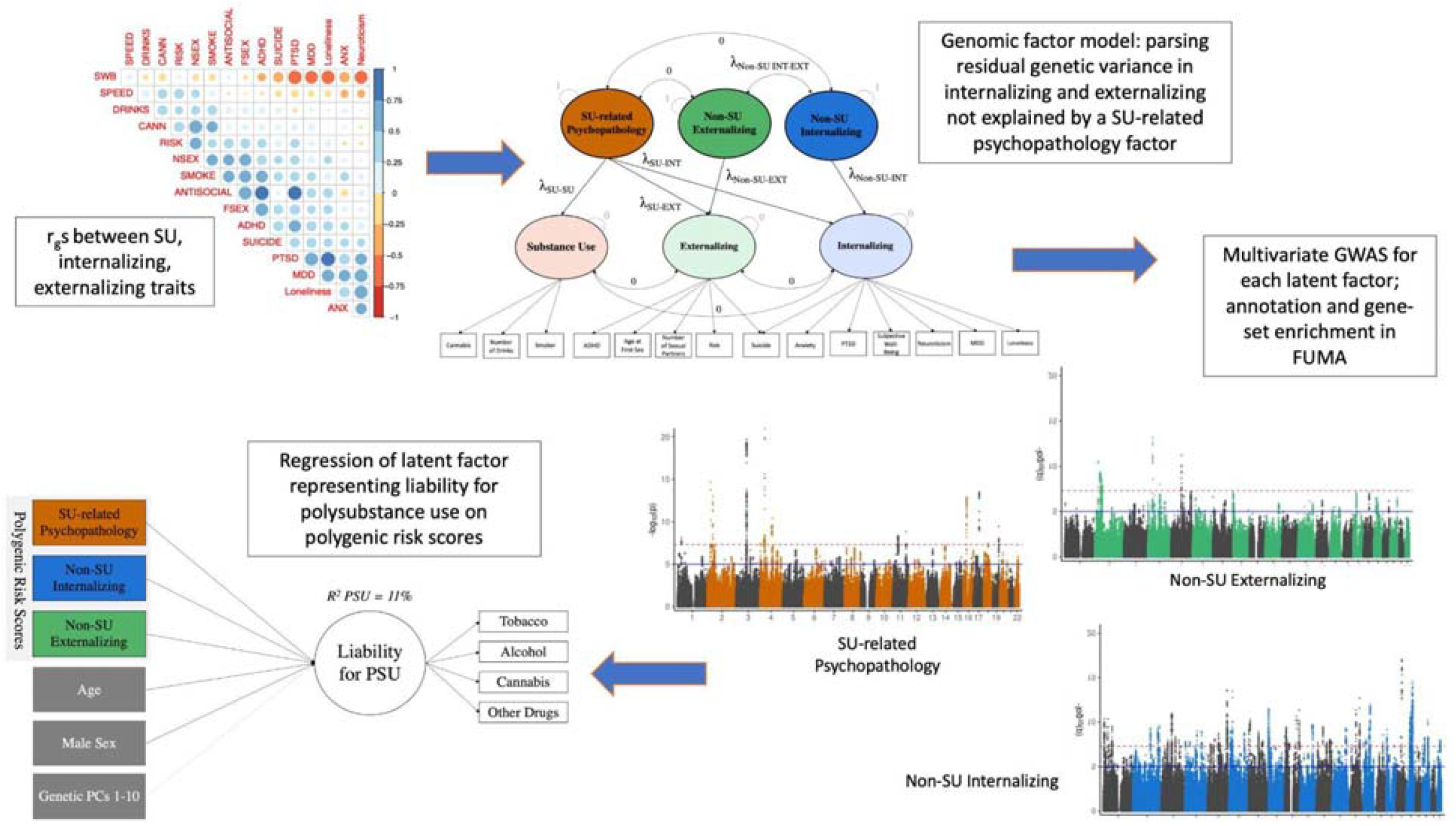
Methods Overview.

### Selection of GWAS summary statistics and target dataset

#### GWAS Summary Statistics

GWAS summary statistics were selected from publicly-available GWAS of individuals of European ancestry on a variety of internalizing, externalizing, and substance use traits. These datasets have been extensively described elsewhere (see original publications), but are summarized in **Table 1**. Studies were selected on the basis of availability of summary statistics, large sample size, and year published, with larger, more recent studies prioritized.

**Table 1.** Summary of GWAS datasets.

| **Domain** | **Description** | **Author (Year)** | **N** |
| --- | --- | --- | --- |
| *Internalizing* | Loneliness | Day et al (2018) | 445,024 |
| *Internalizing* | Depression | Howard et al (2018) | 500,199 |
| *Internalizing* | Neuroticism | Baselmans et al (2019) | 523,783 |
| *Internalizing* | Subjective wellbeing | Okbay et al (2016) | 298,420 |
| *Internalizing* | PTSD | Nievergelt et al (2019) | 174,659 |
| *Internalizing* | Anxiety disorder factor | Otowa et al (2016) | 18,186 |
| *Internalizing/*<br>*Externalizing* | Suicide death | Docherty et al (2020) | 16,873 |
| *Externalizing* | Risk taking behavior | Karlsson Linnér et al (2019) | 1,100,000 |
| *Externalizing* | Number of sexual partners | Karlsson Linnér et al (2019) | 370,711 |
| *Externalizing* | ADHD diagnosis | Demontis et al (2019) | 35,107 |
| *Externalizing* | Age at first sex | Karlsson Linnér et al (2021) | 317,694 |
| *Externalizing* | Antisocial behavior | Tielbeek et al (2017) | 16,400 |
| *Externalizing* | Automobile speeding | Karlsson Linnér (2019) | 404,291 |
| *Substance Use* | Smoking initiation | Liu et al (2019) | 632,802 |
| *Substance Use* | Cannabis use | Pasman et al (2018) | 162,082 |
| *Substance Use* | Drinks per week | Liu et al (2019) | 537,349 |

##### Externalizing Psychopathology and Substance Use

Selection of externalizing psychopathology and substance use traits was partly based on a recent genomic SEM investigation of externalizing (24) and included three measures of substance use: lifetime cannabis use (N=162,082)(25), lifetime smoking initiation (N=632,802)(26), and the number of alcoholic drinks per week (N=537,349)(26). Additionally, we included four measures of non-substance use externalizing problems, including attention-deficit/hyperactivity disorder (ADHD; N=55,290)(27), general risk tolerance and speeding behavior (N=939,908)(24, 28), reverse-coded age at first sexual intercourse (N=317,694) and number of sexual partners (N=370,711) obtained from the UK Biobank (http://www.nealelab.is/uk-biobank/), and antisocial behavior (29).

##### Internalizing Psychopathology

Selection of internalizing psychopathology traits was based on seven recent GWASs. We included loneliness (N=445,024)(30), major depressive disorder (N=500,199)(31), neuroticism (N=523,783)(32), subjective wellbeing (N=298,430)(33), post-traumatic stress disorder (PTSD, N=174,659)(34), a factor score for anxiety disorders (N=18,186)(35), and suicide death (N=16,873)(36).

#### Target Dataset

Target data were drawn from the National Longitudinal Study of Adolescent to Adult Health (Add Health), a nationally representative sample of 20,745 youth starting in grades 7-12 in the United States. Specifically, substance use and demographic data were drawn from the Wave IV survey, which occurred from 2008-2009, on a subset of participants (N=15,071). Participants ranged in age from 24-34 (mean age=28.98 years; SD=1.75). Additional information can be found at http://www.cpc.unc.edu/projects/addhealth/design. All participants in Add Health provided written, informed assent/consent for participation per the University of North Carolina School of Public Health Institutional Review Board guideline: https://www.cpc.unc.edu/projects/addhealth/faqs/index.html#Was-informed-consent-required.

### Construction of Genetic Factor Model

The multivariate genetic model of substance use, internalizing, and externalizing was fitted using the genomic SEM package in R (Grotzinger et al., 2019), which leverages linkage disequilibrium score regression (Bulik-Sullivan et al., 2015) to create a genetic covariance matrix among all traits based on the GWAS summary statistics (see **Supplementary Materials**).

Multivariate genetic models were fitted from the resulting genetic covariance matrix (see **Supplemental Table S1**) and we fitted a series of theoretically plausible confirmatory factor models (see **Supplementary Table S2**). We began with a common factor model in which all traits loaded on a single factor, then tested a correlated two-factor model in which substance use traits and externalizing traits loaded on one factor and internalizing traits loaded on a second factor, followed by a correlated three-factor model in which substance use, externalizing, and internalizing traits loaded on separate factors. Traits were removed from analyses if they resulted in issues related to large differences in Z-statistics due to smoothing of non-positive definite matrices (e.g., due to low powered traits) or did not load highly on a factor (e.g., factor loading less than .3). The best fitting model was selected on the basis of fit, parsimony, and interpretability and used for all subsequent analyses.

Briefly, models indicated that speeding behavior did not load well on the hypothesized factor (externalizing) and it was dropped from the model. When antisocial behavior was included in the model, a difference greater than 0.025 in the genetic covariance matrix was observed pre- and post-smoothing for Z-statistics, therefore it was also dropped. Given that suicide death is sometimes conceptualized as both internalizing and externalizing (37), it was allowed to cross-load on both internalizing and externalizing. The final model was a correlated, three-factor model representing 14 traits across substance use, internalizing, and externalizing domains, (*χ*^2^ (73) = 2597.81, CFI = 0.90, SRMR = 0.11, AIC = 2661.81).

Next, we reparametrized the model using higher order factors consistent with Demange, Malanchini (23) to decompose the variance in internalizing and externalizing components that were shared and not shared with the substance use factor (see **Figure 2 panel a**). This model had identical fit to the prior three-factor model, but the higher order factors facilitate the separation of residual genetic variance in internalizing and externalizing factors not explained by substance use traits. Thus, the three resulting higher-order factors represented: 1) substance use (SU)-related psychopathology (including variance shared across substance use, internalizing, and externalizing psychopathology), 2) variance in internalizing traits not related to substance use (non-SU internalizing), and 3) variance in externalizing traits not related to substance use (non-SU externalizing).

**Figure 2.**
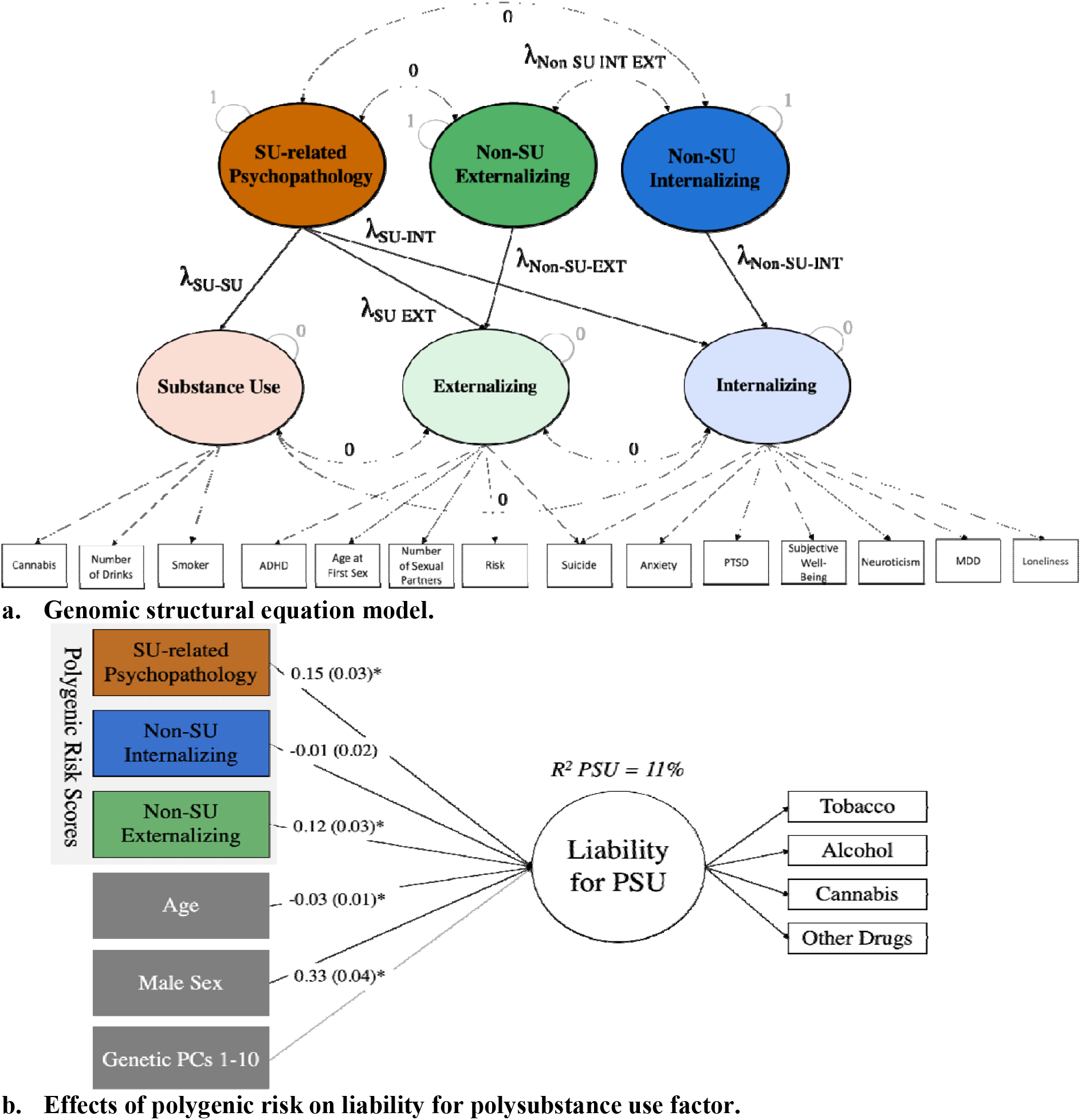
Path Diagrams of Primary Models Tested. **Panel a)** Path diagram of the Genomic Structural Equation Model in which common variance across substance use (SU) related psychopathology is partitioned from common variance in externalizing (EXT) and internalizing (INT) traits using higher order factors. *Note: Numbers along paths indicate the value at which that parameter is fixed. Lambdas (*λ *are estimated*. **Panel b)** Path diagram of the regression of the latent factor indicating liability for polysubstance use (PSU) on polygenic risk scores and covariates, χ (47)=93.61, p<0.001, CFI=0.91, RMSEA=0.02. *Note: For simplicity, parameter estimates not shown for the genetics principal components (PCs) or item factor loadings for the PSU factor. All estimates are unstandardized. SU = substance use. * indicates p < 0*.*05*.

### Multivariate GWAS-by-subtraction

Based on the results from the final genetic factor model, we conducted a GWAS-by-subtraction model to estimate SNP-level associations for each factor. Thus, the multivariate GWAS allowed for three paths of association for each SNP (total # SNPS = 1,558,750): one path mediated by the SU-related psychopathology factor and the other two paths for the non-SU internalizing and non-SU externalizing factors. Subsequently, we performed a Q-analysis which provides an estimate of the extent to which each SNP’s effect is mediated by the factor model rather than the individual items contributing to the factor(s). Larger Q-values indicate more heterogeneity and an increased likelihood that the effect of the SNP is not mediated by the factor, but rather acting directly on an individual trait. Based on the results of the Q-analysis, 1,720 variants were identified as likely to be contributing directly to indicators rather than being mediated by the specified factors. To be conservative, these variants were dropped for subsequent analyses resulting in 1,557,030 SNPs; full GWAS results are presented in **Supplementary Figure S2**.

We calculated the effective n’s for each factor consistent with the approach in Demange, Malanchini (23); these sample sizes are as follows: 1,734,340 (SU psychopathology-related), 1,164,731 (Non-SU internalizing), and 730,198 (Non-SU externalizing). See **Supplementary Materials** for more details.

### FUMA

We used FUMA version 1.4.2 to perform gene mapping and annotation, gene-set analysis, and visualize results of the multivariate GWAS (38). We performed gene mapping using positional mapping (based on ANNOVAR annotations), eQTL mapping (using GTEx V8, CommonMind, and BRAINEAC data), and chromatin interaction mapping. We also performed the MAGMA gene, gene-set, and gene expression analyses (using GTEx V8 and BrainSpan data). See **Supplementary Materials** for more details.

### PGS Calculation

We used PRS-CS, which utilizes Bayesian regression and infers posterior SNP effect sizes under continuous shrinkage (CS) priors (39), to calculate polygenic scores. The external reference panel used to adjust for linkage disequilibrium was the 1000 Genomes EUR population (40) and summary statistics were derived from the multivariate GWASs of the three higher-order factors representing SU-related psychopathology, non-SU internalizing, and non-SU externalizing. Genetic data used for the testing sample were comprised of Add Health participants who were of European genetic ancestry. See **Supplementary Materials** and **Supplementary Table S3** for a summary of QC and genetic ancestry determination for Add Health data. The PLINK1.9 --score function was used to apply regression weights to the genotype for each SNP and generate PGS based on the sum across all SNPs for each individual.

### Polygenic Risk Model Predicting Polysubstance Use

#### Derivation of Liability for Polysubstance Use Behavior Phenotype

Substance use items were drawn from the Add Health Wave IV survey, including those related to having ever used (e.g., “Have you ever had a drink of beer, wine, or liquor more than two or three times?”) and past 30-day use (e.g., “During the past 30 days, on how many days did you use marijuana?”) across several domains. See **Supplementary Table S4** for a description of original items relating to having ever used substances, having used substances in the past 30-days, and their endorsement rates. We collapsed substance use items across four main substance use domains: tobacco, alcohol, cannabis, and other drug use (e.g., prescription drugs and/or other illicit drugs). Items related to 30-day use within each domain were conceptualized as representing a general liability toward PSU.

Mplus (version 8.7)(41) was used to estimate common variance for PSU items using confirmatory factor analysis (CFA). See **Supplementary Materials** for a complete description of the steps taken to build the factor model and **Supplementary Table S5** for endorsement rates of each variable and parameter estimates in the sample (n = 14,000).

CFA indicated that a single factor fit the data well, *χ*^2^ (2) = 6.05, p = 0.049, CFI = 1.00, SRMR = 0.01). Cannabis use loaded highest, followed by other drug use, tobacco use, and alcohol use (range 0.38-0.89). Item thresholds indicated that endorsement of other drug use occurred at the highest values of the latent variable while endorsement of alcohol use occurred at the lowest values of the latent variable. In other words, individuals who reported no use or tended to only use alcohol had the lowest values on the latent trait. In contrast, individuals who endorsed other drug use were among those with the higher values of the latent trait. Broadly speaking, higher scores on the latent trait represented greater liability for PSU while lower scores indicated no use or single substance use.

#### Association of Polygenic Risk Scores with Polysubstance Use Phenotype

We then regressed the PSU factor on the three PGSs (SU-related, non-SU internalizing, non-SU externalizing), controlling for age, sex, and the first 10 genetic principal components using a structural equation modeling approach in Mplus. Due to the complex survey design of the Add Health study, which resulted in an unequal probability of sample selection, clustering by school, and stratification by geographic region, analyses included adjustments using Add Health-provided Wave IV sampling weights, school, and region variables in accordance with Add Health study guidelines (42).

## Results

### Genetic Factor Model of Internalizing, Externalizing, and Substance Use Traits

Genetic correlations among the original 16 traits varied from -0.73 to 0.82 (See **Supplementary Table S1**). The final genomic SEM included three higher order factors to decompose the variance in internalizing and externalizing components that were shared and not shared with the substance use factor. The internalizing and externalizing factors were allowed to correlate. See **Figure 2** (panel a) for a path diagram representing this final model and **Table 2** for a summary of model parameters. The substance use-related psychopathology factor represents common variance across substance use behaviors, internalizing, and externalizing. The non-SU internalizing and non-SU externalizing represents residual variance independent of the SU-related psychopathology shared variance and after accounting for the correlation between internalizing and externalizing.

**Table 2.**
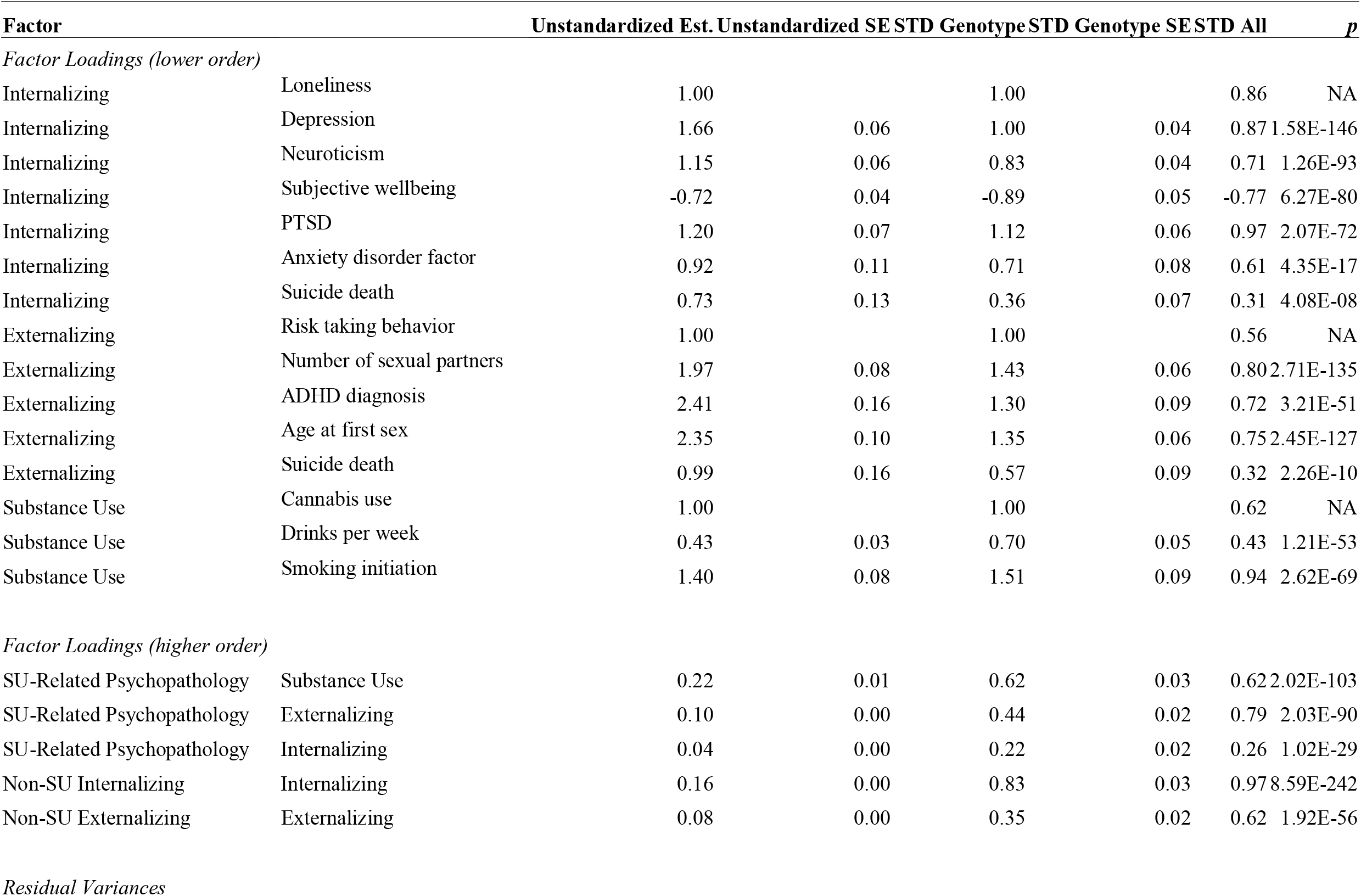
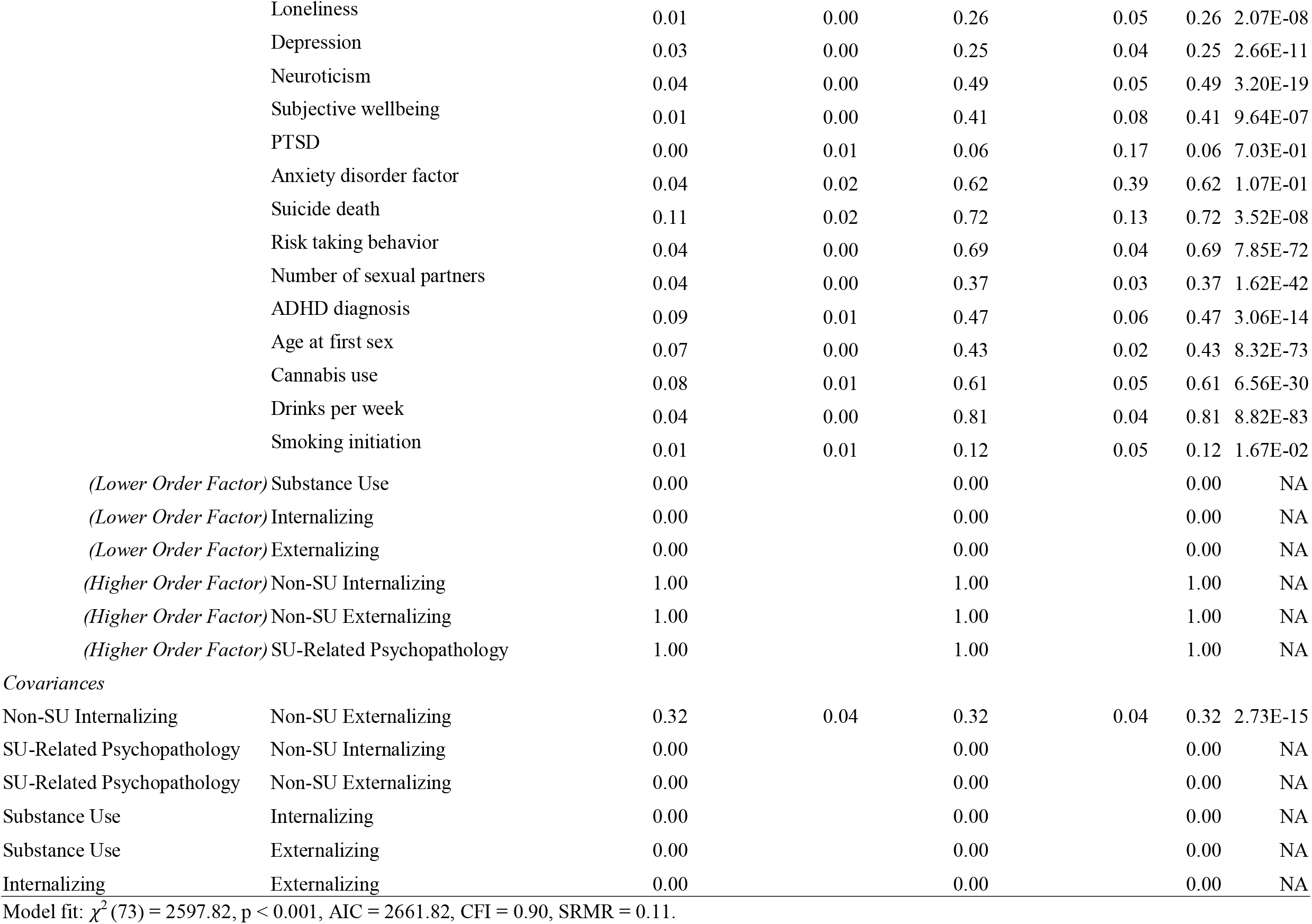
Estimated model parameters for final model representing non-SU internalizing/externalizing factors and SU-related psychopathology factor. *Note: STD Genotype estimates represent estimates that are standardized with respect to variance in traits/phenotypes, STD All estimates are fully standardized, including the endogenous latent variables (i.e., higher-order factors; standard errors are not currently available). SU = substance use, Est = estimate.*

| **Factor** |  | **Unstandardized Est.** | **Unstandardized SE** | **STD Genotype** | **STD Genotype SE** | **STD All** | **p** |
| --- | --- | --- | --- | --- | --- | --- | --- |
| *Factor Loadings (lower order)* |  |  |  |  |  |  |  |
| Internalizing | Loneliness | 1.00 |  | 1.00 |  | 0.86 | NA |
| Internalizing | Depression | 1.66 | 0.06 | 1.00 | 0.04 | 0.87 | 1.58E-146 |
| Internalizing | Neuroticism | 1.15 | 0.06 | 0.83 | 0.04 | 0.71 | 1.26E-93 |
| Internalizing | Subjective wellbeing | -0.72 | 0.04 | -0.89 | 0.05 | -0.77 | 6.27E-80 |
| Internalizing | PTSD | 1.20 | 0.07 | 1.12 | 0.06 | 0.97 | 2.07E-72 |
| Internalizing | Anxiety disorder factor | 0.92 | 0.11 | 0.71 | 0.08 | 0.61 | 4.35E-17 |
| Internalizing | Suicide death | 0.73 | 0.13 | 0.36 | 0.07 | 0.31 | 4.08E-08 |
| Externalizing | Risk taking behavior | 1.00 |  | 1.00 |  | 0.56 | NA |
| Externalizing | Number of sexual partners | 1.97 | 0.08 | 1.43 | 0.06 | 0.80 | 2.71E-135 |
| Externalizing | ADHD diagnosis | 2.41 | 0.16 | 1.30 | 0.09 | 0.72 | 3.21E-51 |
| Externalizing | Age at first sex | 2.35 | 0.10 | 1.35 | 0.06 | 0.75 | 2.45E-127 |
| Externalizing | Suicide death | 0.99 | 0.16 | 0.57 | 0.09 | 0.32 | 2.26E-10 |
| Substance Use | Cannabis use | 1.00 |  | 1.00 |  | 0.62 | NA |
| Substance Use | Drinks per week | 0.43 | 0.03 | 0.70 | 0.05 | 0.43 | 1.21E-53 |
| Substance Use | Smoking initiation | 1.40 | 0.08 | 1.51 | 0.09 | 0.94 | 2.62E-69 |
| *Factor Loadings (higher order)* |  |  |  |  |  |  |  |
| SU-Related Psychopathology | Substance Use | 0.22 | 0.01 | 0.62 | 0.03 | 0.62 | 2.02E-103 |
| SU-Related Psychopathology | Externalizing | 0.10 | 0.00 | 0.44 | 0.02 | 0.79 | 2.03E-90 |
| SU-Related Psychopathology | Internalizing | 0.04 | 0.00 | 0.22 | 0.02 | 0.26 | 1.02E-29 |
| Non-SU Internalizing | Internalizing | 0.16 | 0.00 | 0.83 | 0.03 | 0.97 | 8.59E-242 |
| Non-SU Externalizing | Externalizing | 0.08 | 0.00 | 0.35 | 0.02 | 0.62 | 1.92E-56 |
| *Residual Variances* |  |  |  |  |  |  |  |
|  | Loneliness | 0.01 | 0.00 | 0.26 | 0.05 | 0.26 | 2.07E-08 |
|  | Depression | 0.03 | 0.00 | 0.25 | 0.04 | 0.25 | 2.66E-11 |
|  | Neuroticism | 0.04 | 0.00 | 0.49 | 0.05 | 0.49 | 3.20E-19 |
|  | Subjective wellbeing | 0.01 | 0.00 | 0.41 | 0.08 | 0.41 | 9.64E-07 |
|  | PTSD | 0.00 | 0.01 | 0.06 | 0.17 | 0.06 | 7.03E-01 |
|  | Anxiety disorder factor | 0.04 | 0.02 | 0.62 | 0.39 | 0.62 | 1.07E-01 |
|  | Suicide death | 0.11 | 0.02 | 0.72 | 0.13 | 0.72 | 3.52E-08 |
|  | Risk taking behavior | 0.04 | 0.00 | 0.69 | 0.04 | 0.69 | 7.85E-72 |
|  | Number of sexual partners | 0.04 | 0.00 | 0.37 | 0.03 | 0.37 | 1.62E-42 |
|  | ADHD diagnosis | 0.09 | 0.01 | 0.47 | 0.06 | 0.47 | 3.06E-14 |
|  | Age at first sex | 0.07 | 0.00 | 0.43 | 0.02 | 0.43 | 8.32E-73 |
|  | Cannabis use | 0.08 | 0.01 | 0.61 | 0.05 | 0.61 | 6.56E-30 |
|  | Drinks per week | 0.04 | 0.00 | 0.81 | 0.04 | 0.81 | 8.82E-83 |
|  | Smoking initiation | 0.01 | 0.01 | 0.12 | 0.05 | 0.12 | 1.67E-02 |
|  | *(Lower Order Factor)* Substance Use | 0.00 |  | 0.00 |  | 0.00 | NA |
|  | *(Lower Order Factor)* Internalizing | 0.00 |  | 0.00 |  | 0.00 | NA |
|  | *(Lower Order Factor)* Externalizing | 0.00 |  | 0.00 |  | 0.00 | NA |
|  | *(Higher Order Factor)* Non-SU Internalizing | 1.00 |  | 1.00 |  | 1.00 | NA |
|  | *(Higher Order Factor)* Non-SU Externalizing | 1.00 |  | 1.00 |  | 1.00 | NA |
|  | *(Higher Order Factor)* SU-Related Psychopathology | 1.00 |  | 1.00 |  | 1.00 | NA |
| *Covariances* |  |  |  |  |  |  |  |
| Non-SU Internalizing | Non-SU Externalizing | 0.32 | 0.04 | 0.32 | 0.04 | 0.32 | 2.73E-15 |
| SU-Related Psychopathology | Non-SU Internalizing | 0.00 |  | 0.00 |  | 0.00 | NA |
| SU-Related Psychopathology | Non-SU Externalizing | 0.00 |  | 0.00 |  | 0.00 | NA |
| Substance Use | Internalizing | 0.00 |  | 0.00 |  | 0.00 | NA |
| Substance Use | Externalizing | 0.00 |  | 0.00 |  | 0.00 | NA |
| Internalizing | Externalizing | 0.00 |  | 0.00 |  | 0.00 | NA |

Model fit: $\chi^2(73) = 2597.82$ , $p < 0.001$ , AIC = 2661.82, CFI = 0.90, SRMR = 0.11.

### GWAS-by-Subtraction and Functional Mapping and Annotation

#### Substance use-related psychopathology factor

After performing Q-SNP analysis, 1,720 Q-SNPs were removed, leaving 1,557,030 SNPs for analysis. We identified 17 lead SNPs within 15 genomic risk loci (i.e., merging independent lead SNPs within a 250kb window) in the GWAS of the SU-related psychopathology factor (**Figure 3**). MAGMA gene-based analysis identified 51 gene-wise significant genes (correcting for 15,629 genes), with *CADM2* on chromosome 3 being the strongest association. Other notable mapped genes included well-known alcohol-related genes *GCKR, KLB*, and *ADH1B*. No gene sets passed Bonferroni correction in the MAGMA gene-set analysis. MAGMA gene-property tissue expression analyses implicated the pituitary and several brain tissues (cortex, cerebellum, and nucleus accumbens), as well as early-prenatal and early-mid-prenatal developmental stages of brain tissue.

**Figure 3.**
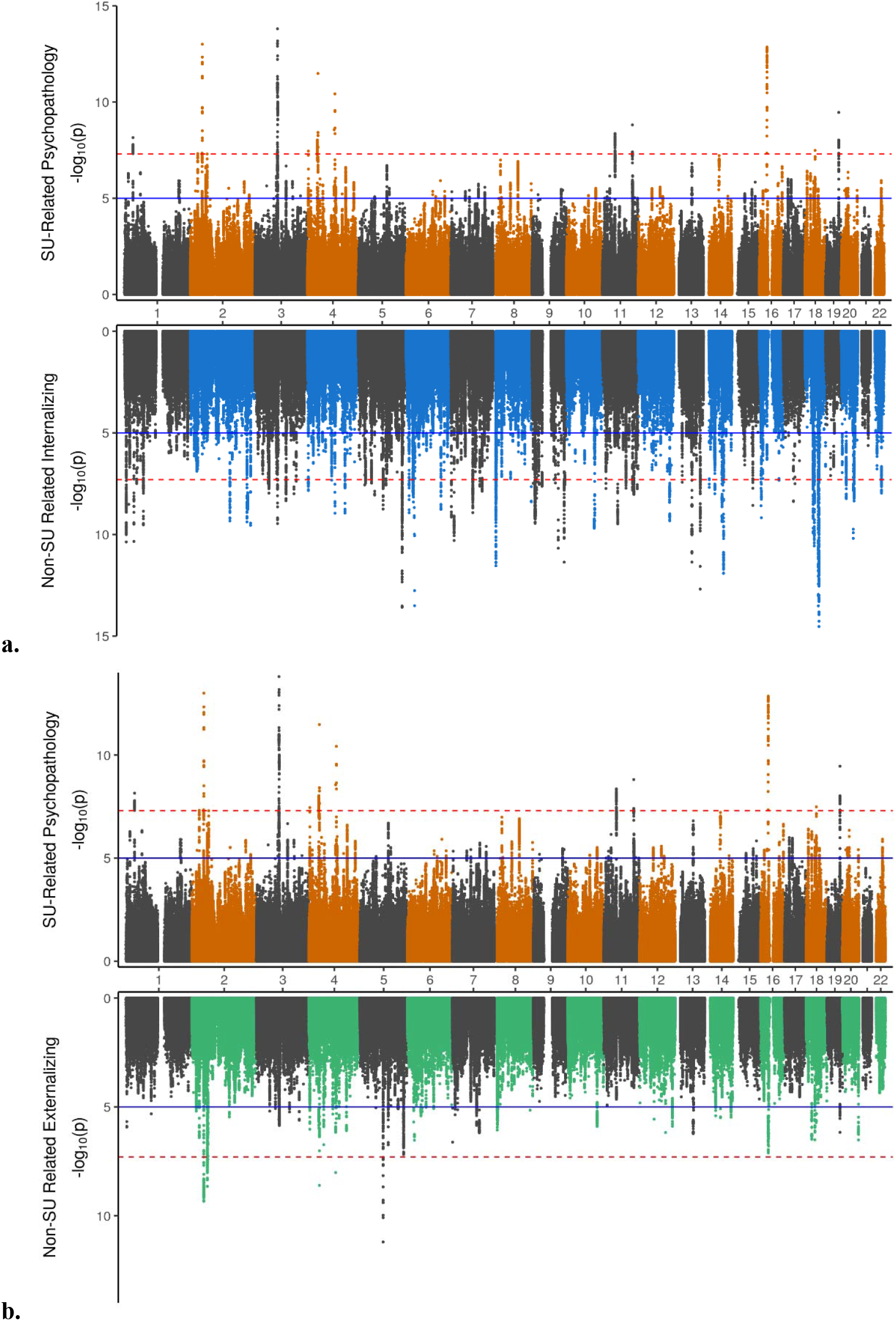
Miami Plots of Multivariate GWAS results. **Panel a)** displays multivariate GWAS results from the Q-SNP-screened sample for substance use-related psychopathology factor (top) and non-substance use internalizing (bottom). **Panel b)** displays multivariate GWAS Q-SNP-screened results for substance use-related psychopathology factor (top) and non-substance use externalizing (bottom).

#### Non-substance use related externalizing factor

For the non-SU externalizing factor multivariate GWAS, we identified 6 genomic risk loci, three on chromosome 2 (lead SNPs rs3931848, rs11682175, and rs7560837), two on chromosome 4 (lead SNPs rs6844176 and rs17028973), and one on chromosome 5 (lead SNP rs4916723; **Figure 3**). Mapped genes included *CAMKMT, VRK2*, and *ADH1B*. MAGMA gene-based analysis identified 27 gene-wise significant genes, some of which overlapped with genes identified via physical mapping for the SU-related psychopathology factor (e.g., *KLB* and *CADM2*). No gene-set associations passed Bonferroni corrections in the MAGMA gene-set analysis. In the MAGMA gene-property tissue expression analyses, pituitary, cortex and cerebellum brain tissues were implicated, as well as early-mid-prenatal developmental stage.

#### Non-substance use related internalizing factor

There were 91 lead SNPs within 80 genomic risk loci identified for the non-SU internalizing factor multivariate GWAS (**Figure 3**). MAGMA gene-based analyses identified 161 genes (top gene: *RBFOX1*), and three gene sets were significant after Bonferroni corrections: one related to inhibin binding (GO_mf:go_inhibin_binding), and two related to neuron differentiation and development (GO_bp:go_neuron_differentiation and GO_bp:go_neuron_development). Other notable mapped genes included *CADM2* and *DRD2*. In MAGMA gene-property tissue expression analyses, all brain tissues except the substantia nigra and spinal cord were significant; the early-mid-prenatal developmental stage of brain tissue was also implicated.

See **Supplementary Tables S6-14** for lists of significant genomic loci, MAGMA gene-based results, and MAGMA gene-set results for each factor.

### Polygenic Score Prediction of Polysubstance Use in Add Health

Our target sample was a large, longitudinal epidemiological study of youth in the United States (the Add Health Study). Among participants who provided data in Wave IV (N=15,701) and were between the ages of 24 and 35 (mean age = 29.0 years, standard deviation = 1.75 years), only 10% reported having never used any substance in their life. The most commonly endorsed substance in the past 30 days was alcohol (60% endorsed), followed by cigarettes (36%), and then cannabis (16%). One third of the sample (34.71%) reported having used more than one substance in the past thirty days, 39.65% reported having used only one substance in the past 30 days, and 25.64% reported having not used any substances in the past thirty days. The most commonly reported combinations of substances in the past 30 days were alcohol and tobacco (16.81%), followed by alcohol, tobacco, and cannabis (6.4%), with lower numbers of people reporting alcohol and cannabis (3.3%) or tobacco and cannabis (2.22%). A single factor solution supported evidence for a general liability for PSU with higher scores representing greater liability for using multiple substances in the past 30 days (see **Supplementary Table S5)**.

Results of the structural equation model in which the latent PSU factor was regressed on the three PGSs (N=4,102) derived from the multivariate GWAS-by-subtraction indicated that PGSs for SU-related psychopathology and non-SU externalizing traits were associated with higher PSU factor scores, *χ*^2^(47)=93.61, p<0.001, CFI=0.91, RMSEA=0.02. The PGS, however, for non-SU internalizing traits was not significantly associated with PSU. Older age was associated with lower risk for PSU and being male was associated with higher risk for PSU. In total, the three PGSs accounted for an additional 4% of the variance in PSU above and beyond a model with only age, sex, and 10 genetic principal components (R^2^ full model = 0.11, R^2^ model without PGSs = 0.07; see **Table 3**).

**Table 3.** Standardized estimates and standard errors (SE) of model estimating polygenic score associations with past 30-day polysubstance use factor (N=4102).

| Model Parameter | Variable | Est | SE | p |
| --- | --- | --- | --- | --- |
| Factor Loadings | Tobacco | 0.63 | 0.04 | <0.001 |
|  | Alcohol | 0.29 | 0.03 | <0.001 |
|  | Cannabis | 0.81 | 0.04 | <0.001 |
|  | Other Drugs | 0.66 | 0.04 | <0.001 |
| Regression Estimates | Substance Use Related Psychopathology | 0.24 | 0.04 | <0.001 |
|  | Non-Substance Use Internalizing | -0.01 | 0.03 | 0.667 |
|  | Non-Substance Use Externalizing | 0.19 | 0.04 | <0.001 |
|  | Age | -0.08 | 0.03 | 0.007 |
|  | Male | 0.26 | 0.03 | <0.001 |
|  | PC1 | 0.03 | 0.03 | 0.362 |
|  | PC2 | -0.03 | 0.03 | 0.296 |
|  | PC3 | -0.04 | 0.04 | 0.312 |
|  | PC4 | <0.01 | 0.03 | 0.935 |
|  | PC5 | 0.03 | 0.04 | 0.406 |
|  | PC6 | -0.07 | 0.04 | 0.116 |
|  | PC7 | 0.02 | 0.05 | 0.687 |
|  | PC8 | <0.01 | 0.05 | 0.939 |
|  | PC9 | -0.06 | 0.03 | 0.026 |
|  | PC10 | 0.04 | 0.04 | 0.324 |
| Item Thresholds | Tobacco | -0.28 | 0.61 | 0.645 |
|  | Alcohol | -1.38 | 0.67 | 0.04 |
|  | Cannabis | 0.14 | 0.64 | 0.822 |
|  | Other Drugs | 0.33 | 0.71 | 0.645 |
| Residual Variance | Polysubstance Use Factor | 0.9 | 0.02 | <0.001 |

Model fit: $\chi^2(23) = 93.61$ , $p < 0.001$ , CFI = 0.91, RMSEA = 0.02

## Discussion

The goals of this study were to distinguish shared and unique genetic variation related to internalizing and externalizing traits that is distinct from common variance related to substance use. We then evaluated whether PGSs derived from these genetic factors independently account for variation in liability for PSU in a national sample of young adults. Thus, using genetic variance from summary statistics across numerous behavioral and psychological traits, we identified three genetic factors that encompassed substance use (cannabis use, cigarettes, and drinking), internalizing (loneliness, depression, neuroticism, subjective wellbeing, PTSD, anxiety, and suicide death), and externalizing (risk taking, number of sexual partners, ADHD, age at first sex, and suicide death) domains. Consistent with the body of literature linking substance use behavior with both internalizing and externalizing traits, we used the GWAS-by-subtraction approach (23) to reparametrize the model into three higher-order factors representing: 1) genetic variance shared across substance use, internalizing, and externalizing traits (i.e., SU-related psychopathology), 2) variance in internalizing traits not related to substance use (non-SU internalizing), and 3) variance in externalizing traits not related to substance use (non-SU externalizing).

Next, we conducted a multivariate GWAS to examine the genetic underpinnings of the three factors. After excluding significant Q-SNPs (i.e., SNPs which are not mediated by the latent factor) from each of the factor results, we identified 15 genomic risk loci for the SU-related psychopathology factor, 80 risk loci for the non-SU internalizing factor, and 6 risk loci for the non-SU externalizing factor. Interestingly, the strongest gene-based finding for the SU-related psychopathology factor was *CADM2* (**Supplementary Table S9**), which has been linked to many different behavioral traits and psychiatric disorders (43). This gene was also mapped to risk loci in the non-SU internalizing factor and the non-SU externalizing factor, consistent with prior studies suggesting that this gene might be a shared risk factor for many related behavioral and psychiatric phenotypes. Perhaps unsurprisingly, we found far fewer genetic risk loci for non-SU externalizing (6 loci) than for non-SU internalizing (80 loci). This could be due to more GWASs loading on the internalizing factor than the externalizing factor, and/or greater shared variance between SU and externalizing than for SU and internalizing. It is worth noting that we did not have access to 23andMe summary statistics; thus, it is difficult to make comparisons to the largest prior effort to study the genetics of externalizing (24), which included some GWAS meta-analyses with 23andMe data and thus had greater statistical power than the current multivariate GWAS.

All three factor GWASs showed enrichment in a number of brain tissues, and the SU-related and non-SU internalizing factors were also enriched for pituitary tissues (**Supplementary Figure S3**). Each GWAS was enriched for early-mid-prenatal stages of brain development, and the SU-related psychopathology GWAS was additionally enriched for the early-prenatal stage of development. Only the non-SU internalizing factor GWAS revealed significant associated gene-sets related to neuron differentiation and development and inhibin binding, but this may be more of a reflection of statistical power than true differences in gene-set associations compared to the other two GWASs, as many of the top 10 gene-sets overlapped between the non-SU internalizing and non-SU externalizing GWAS (e.g., neuron development, neurogenesis; see **Supplementary Tables S13 and S14**.

Finally, we constructed PGSs for each factor using weights from the multivariate GWAS to evaluate whether genetic variance for each domain were associated with liability for PSU in the Add Health Study. We parameterized PSU as a common factor underlying liability for endorsing past 30-day use of tobacco, alcohol, cannabis, or other illicit drugs. After regressing the PGSs on the PSU factor, our findings indicated that the SU-related psychopathology PGS was associated with greater liability for PSU. More importantly, our findings also indicated that the PGS representing non-SU externalizing traits (i.e., residual genetic variance in externalizing that was separate from substance use related psychopathology) also contributed to greater liability for PSU, above and beyond the SU-related psychopathology score. This was not true for the non-SU internalizing traits, however. In other words, even after accounting for the genetic variance in externalizing that is shared with substance use, PGSs of non-SU externalizing traits were still associated with higher liability for PSU.

Interestingly, these findings suggest that there may be unique genetic variance in externalizing traits contributing to liability for PSU that is independent of the genetic variance shared with SU. It is important to note that our model partitioned common variance based on GWAS summary statistics that were available at the time of writing; thus, these results may look different if we were able to include genetic variance from other substances or other behaviors.

For example, we included alcohol, cannabis, and smoking behaviors in our model but were unable to include other substances due to the lack of large GWAS summary data available for other (especially illicit) substances like cocaine, opioids, or hallucinogens. Thus, the common and unique variance of the higher order externalizing factor may look different had we included other traits. For example, behaviors like sensation seeking, impulsivity, and experimentation might be highly relevant to PSU, but are not captured in the present SU-related factor and therefore contribute to the non-SU externalizing factor in our model. A further explanation for the additional genetic variance in non-SU externalizing could be that this variance is showing up as a distinct factor because of the very nature by which this construct was designed. For example, individual differences in the ability to self-regulate one’s behavior in a non-substance context is also a hallmark of antisocial behaviors. As more GWASs of substance use and externalizing traits become available, future work could expand on these findings to examine whether results are consistent when a broader set of substance use traits are integrated, including whether SUD or lifetime ever use is considered.

The current study highlights the utility of GWAS to identify and characterize mechanisms that are general and specific to multiple psychological traits. However, this study should be interpreted in light of several limitations related to our understanding of the genetic epidemiology of the selected behaviors. The construction of genomic SEMs from summary statistics data requires large sample sizes for robust models. Consequently, we only included traits for which large, publicly available GWAS summary statistics could be obtained. For example, while we intended to include a GWAS of antisocial behavior (29), inclusion of this data resulted in model convergence issues and we had to exclude it.

While the focus of this work was on substance use, research suggests that the etiology of substance use phenotypes are only partially related to the mechanisms that drive individual differences in substance *dependence/use disorder*. As highlighted in Gelernter and Polimanti (22), relative to the use-based traits, SUD traits tend to have higher genetic correlations with psychopathology. Indeed, large scale GWASs examining alcohol-related phenotypes have found distinct patterns of genetic correlations between alcohol consumption and alcohol problems (44). Associations between dependence and problematic use with psychiatric traits have been consistently found in subsequent studies, suggesting that genetic risk factors related to problems, rather than consumption or use behavior, distinctly contribute to shared vulnerability for psychiatric disorders (22). For example, a large GWAS of cannabis use disorder evidenced moderate genetic correlation between CUD and cannabis use initiation (r_g_=0.50), while another study found that repeated cannabis use, but not cannabis use initiation, was moderately genetically associated with an index of traits related to vulnerability to substance use disorder (r_g_=0.76)(45). Thus, as larger GWASs of substance dependence phenotypes become available, future work could seek to examine distinctions between how genetic risk for substance use versus dependence confer risk for polysubstance use behavior.

Furthermore, while the PSU phenotype may represent a potentially distinct, or perhaps more severe SU phenotype typology from lifetime ever-use of multiple substances, this conceptualization is unable to disentangle potential nuances related to co-use (i.e., concurrent use) of specific substances together. For example, individuals who co-use substances are at great risk for adverse health reactions and may be genetically distinct from casual or experimental users. Distinguishing these nuanced effects will be important for future research as individuals with greater liability for using multiple substances may have different disease courses, putting them at risk for SUD, or may possess a greater degree of genetic variance related to maladaptive traits.

Finally, these results must also be interpreted in light of the sample used, which consisted of young adults of European ancestry from a national epidemiological study in the US. To date, there are few GWASs of behavioral traits, particularly in the externalizing domain, conducted among individuals from other ancestral groups. Consequently, at this time, we are unable to examine genetic correlations between substance use and internalizing/externalizing domains or create PGSs that are specific to other ancestral groups due to the lack of representation of GWAs across multiple ancestries, moreover in similar contextual environments as the United States. As more diverse GWASs become available, the results from this work should be repeated among samples that differ with regards to ancestry, nationality, and age.

In summary, GWAS across multiple psychiatric disorders reflect a combination of shared and unique genetic effects that can inform individual differences in PSU. Adjudication of polygenic liability, inferred from large GWAS meta-analyses, using factor analytic approaches will provide much needed insight into how to interpret and leverage polygenic scores in future endeavors.

## Data Availability

Data used in this study is publicly available. Summary statistics for presented analyses available upon request.

