## Supplementary Materials for "Genetic associations among internalizing and externalizing traits with polysubstance use among young adults"

**Preparation and Analysis of Base Data: GWAS Summary Statistics**

**Genetic Structural Equation Models.** The multivariate genetic model of substance use, internalizing, and externalizing was fitted using the GenomicSEM package in R (1). Genomic SEM leverages linkage disequilibrium score regression (LDSC)(2) to create a genetic covariance matrix among all traits based on the GWAS summary statistics. Adjustments for sample overlap among GWAS are made by estimating a sampling covariance matrix that indexes the extent to which the sampling errors of the estimates are associated with one another Grotzinger (1).

GWAS summary statistics files were munged using the GenomicSEM version 0.0.5c. Markers with MAF < .01 were screened out prior to running LDSC and obtaining the genetic variance/covariance matrix. See **Supplementary** **Table S1** for genetic correlations among all traits used in the current study. Next, multivariate genetic models were fitted using the genetic covariance matrix as input in the same way that traditional SEM models are fitted to a covariance matrix, drawing on functionality from the lavaan R package Rosseel (3) and used the default diagonally weighted least squares estimation method. Model fit was determined using chi-square tests (χ^2^), the Comparative Fit Index (CFI), and the Standardized Root Mean Square Residual (SRMR). Models with good fit typically have CFI values greater than .95 and SRMR values lower than .08. Additionally, although good-fitting models also traditionally have nonsignificant χ^2^ statistics, we did not use this criterion as GWASs are extremely large and this statistic is sensitive to sample size. Given that studies using genomic SEM based on these large-scale GWASs have generally used more relaxed thresholds for other fit statistics such as CFI >.90 (4); we adopted this threshold for acceptable fit in the current study.

**Parameterization of the final model using GWAS-by-subtraction.** To fit a higher-order model in which residual variance for internalizing and externalizing factors were fit separate from common variance shared with substance use behaviors. In this model, we included three higher-order factors in the final, three-factor model with substance use, internalizing, and externalizing factors. The higher order substance use related psychopathology factor was regressed on all three lower-level factors (substance use, internalizing, externalizing). The higher order non-substance use externalizing factor was regressed on the lower-order externalizing factor and the higher order non-substance use internalizing factor was regressed on the lower order internalizing factor. These two factors represent the residual genetic variance in internalizing and externalizing domains after variance related to substance use was regressed out. Each higher order factor variance was fixed to 1 and each lower order factor variance was fixed to 0. Covariances between all lower order factors were fixed to 0. The covariance between non-substance use internalizing and non-substance use externalizing was allowed to vary, while the covariance between substance use-related psychopathology and the other two higher order factors (non-substance use internalizing, non-substance use externalizing) were fixed to 0. **See Figure 2.** For the multivariate GWAS, each of these three higher order factors was regressed on SNPs.

**Alternate Results: Full GWAS Data (including Q-SNPs).** Our primary analyses screened out markers that were identified to have high heterogeneity. Please see **Supplemental Figure S2** for results containing the full set of SNPs from the multivariate GWAS.

**Effective N calculation.** To calculate an effective N (Neff) for our multivariate GWAS, a lower and upper bound for the Minor Allele Frequency (MAF) was set at 10% and 40% respectively to reduce error introduced from SNPs with low MAF. Following the calculations laid out by Demange and colleagues (2021), SNP effect estimates were multiplied by the residual heritability estimates of each factor. Through this, we estimated effective n’s of 1,734,340 (SU-related), 1,164,731 (Non-SU internalizing), and 730,198 (Non-SU externalizing) for each of the factors.

**FUMA.** We used FUMA version 1.4.2 to perform gene mapping and annotation, gene-set analysis, and visualize results of the multivariate GWAS Wantanabe (5). After filtering out SNPs with significant heterogeneity as evidenced by a Q-SNP p<5e-8, we loaded the summary statistics for each factor onto the FUMA web-based platform. We used the effective N (calculated as described above) for each factor as our sample size parameter in FUMA.

We defined “lead SNPs” as those which reached genome-wide significance (p < 5e-8) and were independent of each other at an r^2^ < 0.1. “Genomic risk loci” were defined by merging LD blocks of lead SNPs within a 250 kb distance; that is, if there existed two lead SNPs (and their associated blocks of SNPs in high LD with the lead SNP) within 250 kb distance of each other, even if the lead SNPs are independent at r^2^ < 0.1, they would be defined by a single genomic risk locus.

We performed gene mapping using positional mapping (based on ANNOVAR annotations), eQTL mapping (using GTEx V8, CommonMind, and BRAINEAC data), and chromatin interaction mapping. We also performed the MAGMA gene, gene-set, and gene expression analyses (using GTEx V8 and BrainSpan data).

See **Supplementary Tables S6-14** for lists of significant genomic loci, MAGMA gene-based results, and MAGMA gene-set results for each factor.

**Preparation and Analysis of Target Data: Add Health**

**Ancestry Determination.** To identify genetic ancestry, we conducted principal components analysis (PCA) anchored with the 1000 Genomes Project (1KG) Phase III (Version 5) reference panel (6) using FlashPCA2 (7). The goal of this step was to obtain a homogenous group of individuals based on empirically determined, genetically informed ancestral groups anchored by a widely used reference panel containing several super-populations (i.e., African, Ad Mixed American, East Asian, European, and South Asian). First, markers with genotyping rate >95% and MAF >10% were selected and aligned to the 1KG data (i.e., strand alignment and allele frequencies compared to European Ancestery1KG reference panel)(8). Next, individuals who failed a multidimensional outlier test were considered outliers and excluded. The first step of this test involved identifying and excluding individuals whose principal component values fell beyond two standard deviations from the first three principal component means derived from the members of each super-population (see **Supplementary Figure S4**). Then, multidimensional distance scores for each individual were calculated and distance scores were compared to a sample-level outlier threshold based on the interquartile range. A total of 5,437 individuals of European Ancestry (EA; the largest homogenous population) were identified and selected from the data for imputation.

**Genotype Imputation.** Following ancestry determination and to prepare the data for imputation, markers with genotyping rate >95%, MAF >1%, individuals with missingness >90% were selected and aligned to the Haplotype Reference Consortium (HRC) reference data. Next, the sample was genetically imputed using the HRC r1.1 2016 reference panel and Eagle v2.4 phasing with Minimac4 via the Michigan Imputation Server (https://imputationserver.sph.umich.edu/index.html#!pages/home). Following imputation, markers that were not bi-allelic, were not autosomal, or had poor imputation quality score (*r2* < 0.70) were removed. Next, markers that had a call rate < 99%, low minor allele frequency (< 1%), or failed HWE test (*p* < 0.0001) were removed and samples with < 90% missing data were removed, resulting in a total of 9,735,354 SNPs (see **Supplementary Table S3** for an outline of all QC and imputation steps). Finally, a genetic relationship matrix (GRM) was computed using the GCTA software tool [version 1.25.3]. A total of 4,726 unrelated (relatedness <5%) individuals of EA and 6,671,511 SNPs were retained for analysis.

**Derivation of PSU Factor Scores.** Derivation of PSU factor scores were conducted in a stepwise manner using R. Data were gathered from Wave 4 of the Add Health study. Past 30-day substance use variables were calculated for the following substances: tobacco, alcohol, cannabis, and other drugs (i.e., prescription drugs and other illicit drugs) and coded as 0/1 (endorsed past 30-day use or not endorsed). See **Supplementary Table S4** for a table summarizing substance use items and their endorsement rates in the Add Health sample. After item selection, data were randomly subset into two samples for Principal Components Analysis (PCA; n = 7851) and Confirmatory Factor Analysis (CFA; n = 7850) based on a 50/50 split using the RSample package. PCAs were conducted based on the polychoric correlations between binary 30-day use variables to evaluate whether items loaded on a single component. The number of components for extraction was guided by parallel analysis, which supported a one component model (see **Supplementary Figure S5** for scree plot from parallel analysis).

Following the PCA, a single factor CFA was run on the held-out sample using weighted least squares mean variance estimation (WLSMV) was used given the binary nature of the substance use items. Given that the Add Health study design resulted in an unequal probability of sample selection, analyses accounted for clustering by school, and stratification by geographic region, as well as adjustments using study-provided sampling weights in accordance with Add Health study guidelines (9). **Supplementary Table S5** presents endorsement of each variable in the general sample (N = 15,701) as well as the standardized estimates for factor loading and item thresholds in the CFA sample (n = 14,000 due to missing sampling weights for some participants). Results revealed that this single factor model was a good fit to the data, RMSEA = 0.02, CFI = 1.00, TLI = 0.99. Following construction of the CFA in the held-out sample, a final CFA model was constructed using the entire Add Health Wave 4 sample. This model was a good fit to the data, $\chi$^2^ (2) = 6.05, *p* = 0.049, CFI = 0.998, TLI = 0.993, RMSEA = 0.012, 90% CI [0.001, 0.023]. PSU factor scores from this final CFA model were used in subsequent analyses.

**Supplementary Tables**

*Supplementary tables are located in an Excel file.*

Supplementary Table S1: Genetic correlations.

Supplementary Table S2: Summary of Genomic SEM testing.

Supplementary Table S3: Sample size and number of markers across Add Health data processing and imputation steps.

Supplementary Table S4: Drug Use Variables in Add Health Study (N = 15,701)

Supplementary Table S5: Polysubstance Use Confirmatory Factor Model Results (standardized estimates and standard errors [SE])

Supplementary Table S6: Genomic Risk loci for SU factor

Supplementary Table S7: Genomic Risk loci for non-SU EXT factor

Supplementary Table S8: Genomic Risk loci for non-SU INT factor

Supplementary Table S9: MAGMA gene-based results for SU factor

Supplementary Table S10: MAGMA gene-based results for non-SU EXT factor

Supplementary Table S11: MAGMA gene-based results for non-SU INT factor

Supplementary Table S12: MAGMA gene-set results for SU factor

Supplementary Table S13: MAGMA gene-set results for non-SU Ext factor

Supplementary Table S14: MAGMA gene-set results for non-SU Int factor

**Supplementary Figures**

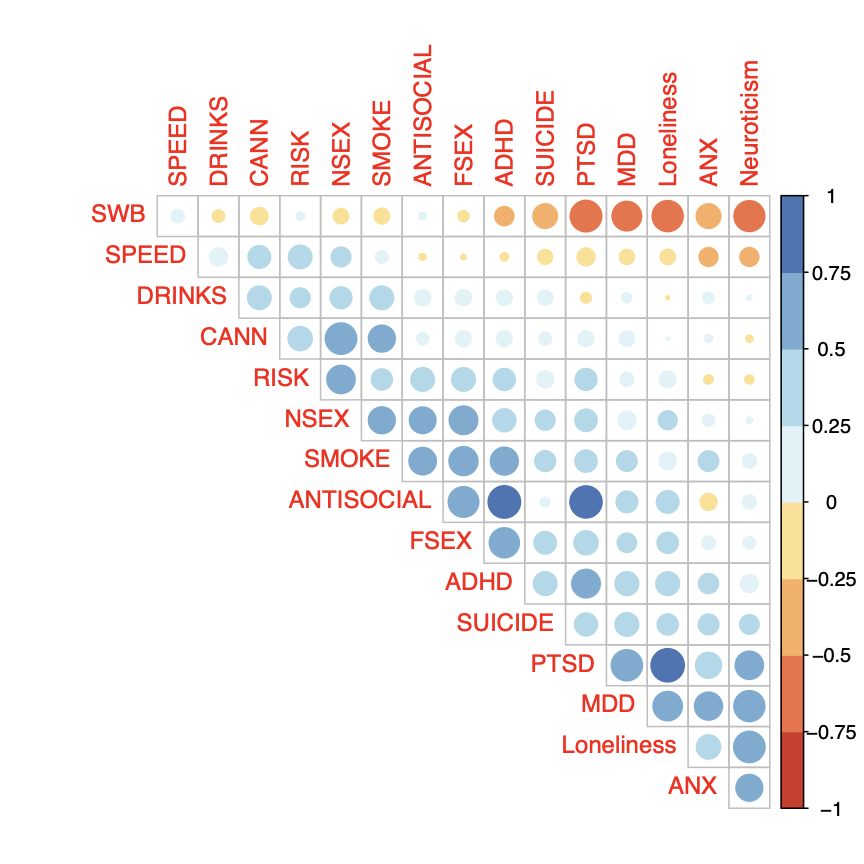
**Supplementary Figure S1.** Heat plot representing correlations among traits from GWAS summary statistics.

*Note: SWB = subjective well-being, SPEED = speeding behavior, DRINKS = number of drinks per week, CANN = cannabis use, RISK = risk tolerance, NSEX = number of sexual partners, SMOKE = smoking (tobacco) initiation, ANTISOCIAL = antisocial behavior, FSEX = age at first sex, ADHD = attention deficit/hyperactivity disorder, PTSD = posttraumatic stress disorder, MDD = major depressive disorder, ANX = anxiety disorder factor score*

**a.**
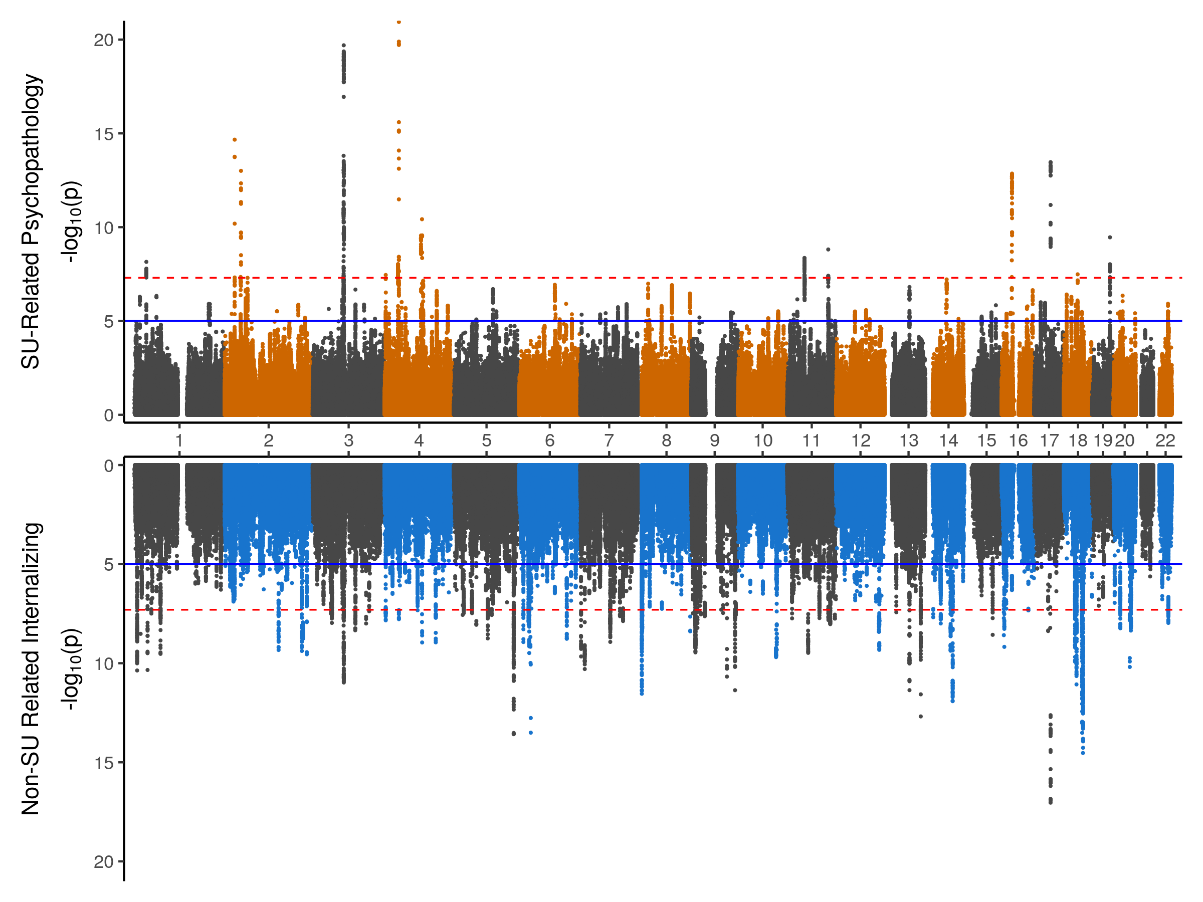

**b.**
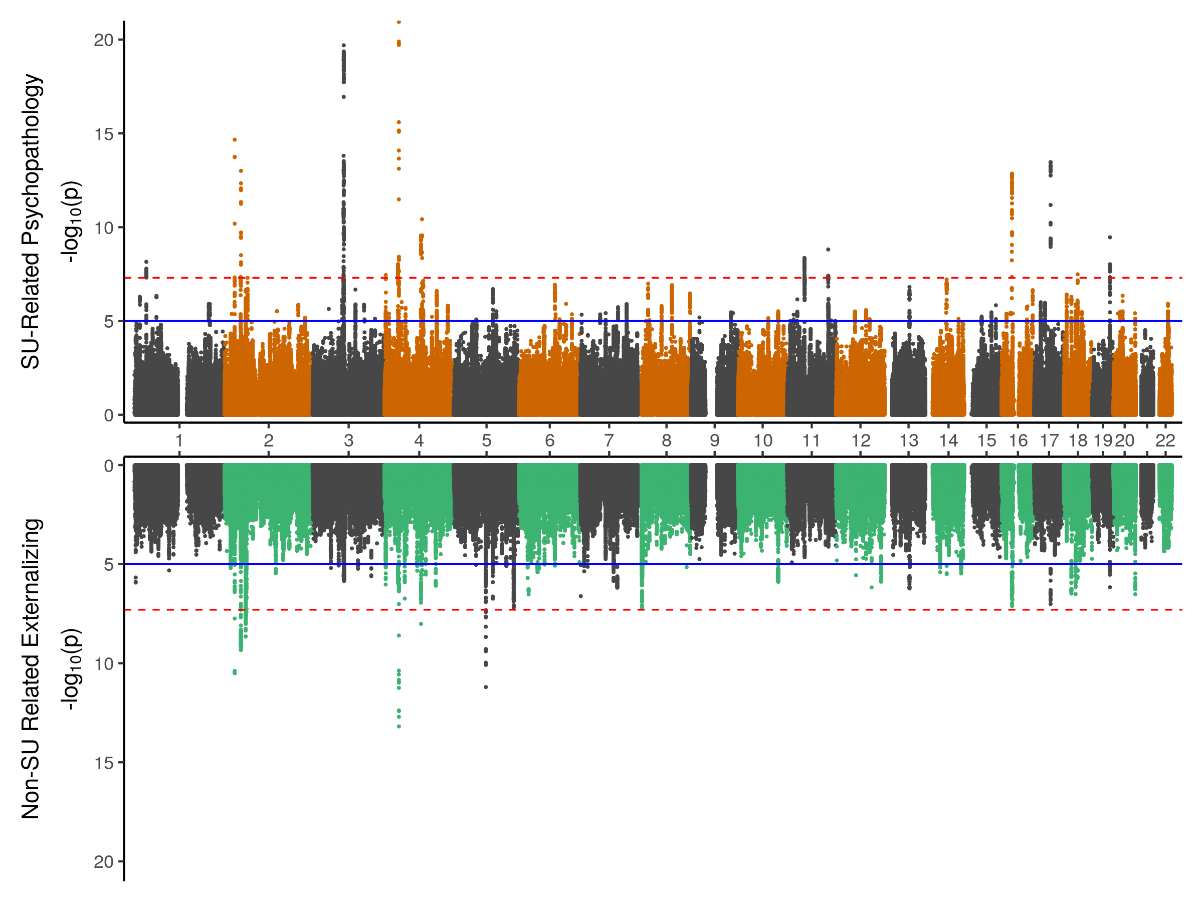

**Supplementary Figure 2. Miami Plots of Multivariate GWAS results using full sample. Panel** **a)** displays the full multivariate GWAS results for substance use-related psychopathology factor (top) and non-substance use internalizing (bottom). **Panel** **b)** displays the full multivariate GWAS results for substance use-related psychopathology factor (top) and non-substance use externalizing (bottom).

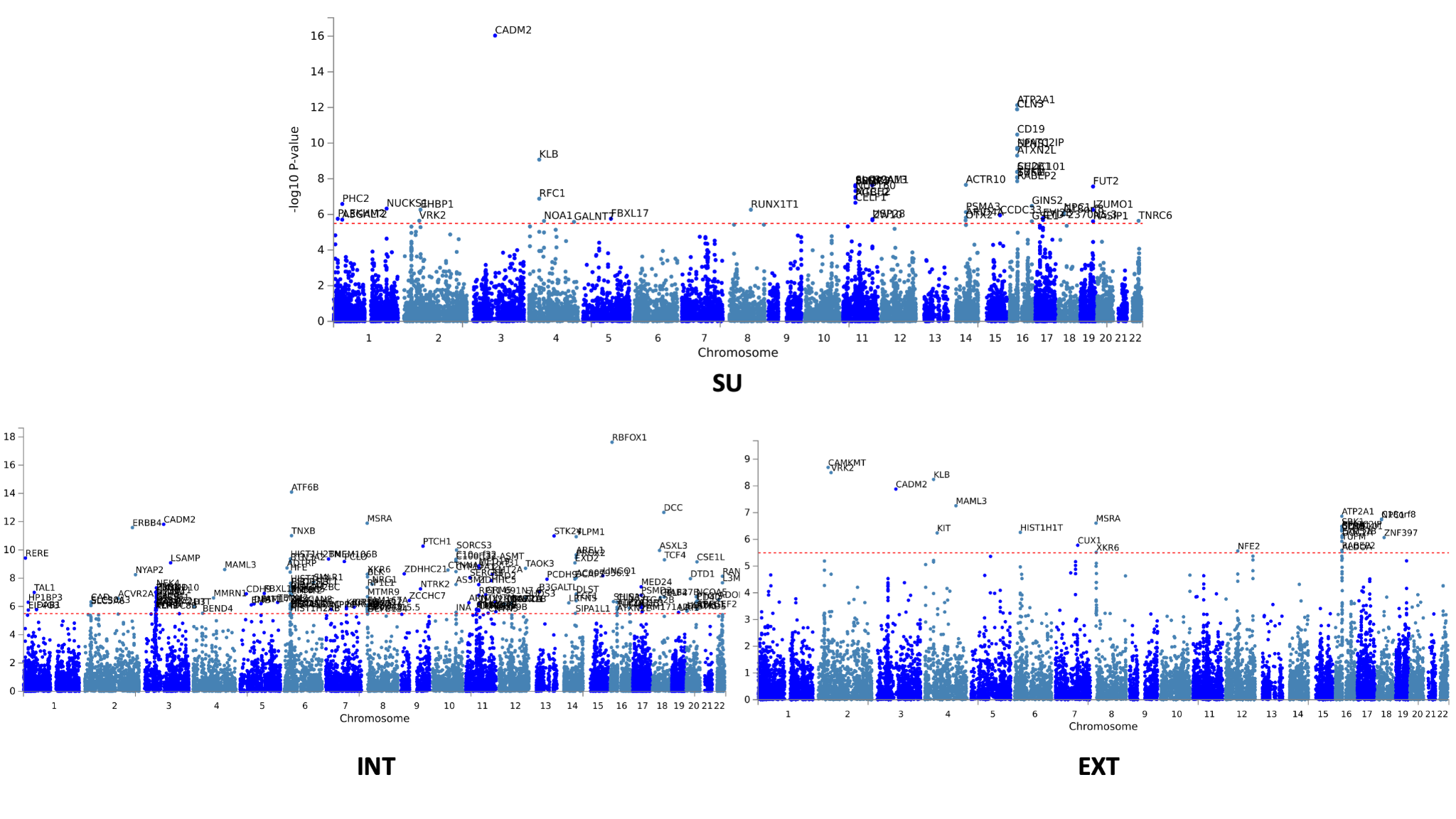

a. **Gene-based Manhattan plots.** There were 51 genes significant for the SU-related factor, 161 significant for the non-SU internalizing factor, and 27 genes significant for the non-SU externalizing factor.

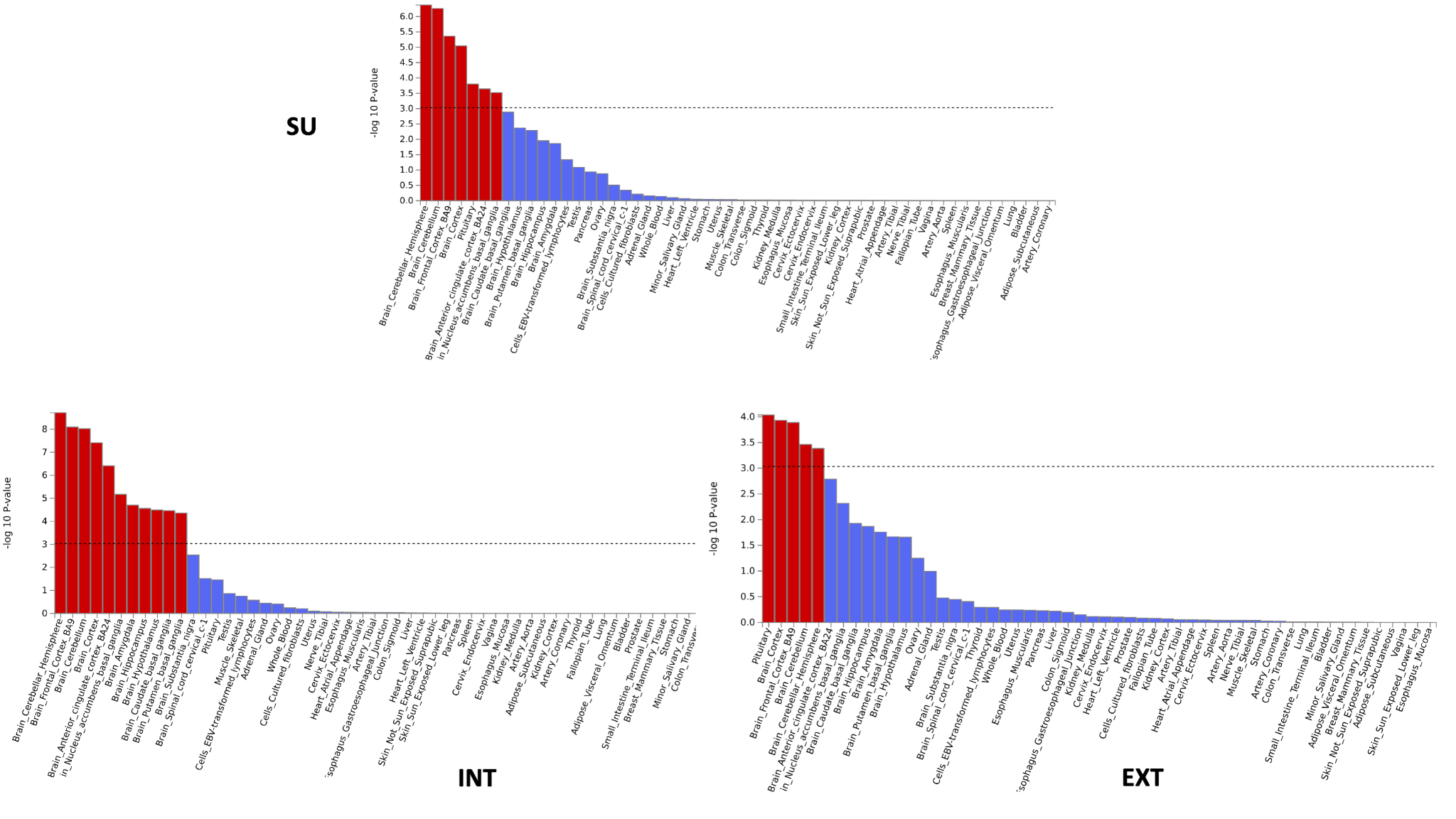

b. **Results from MAGMA tissue expression analysis using GTEx v8 53 tissue types.**

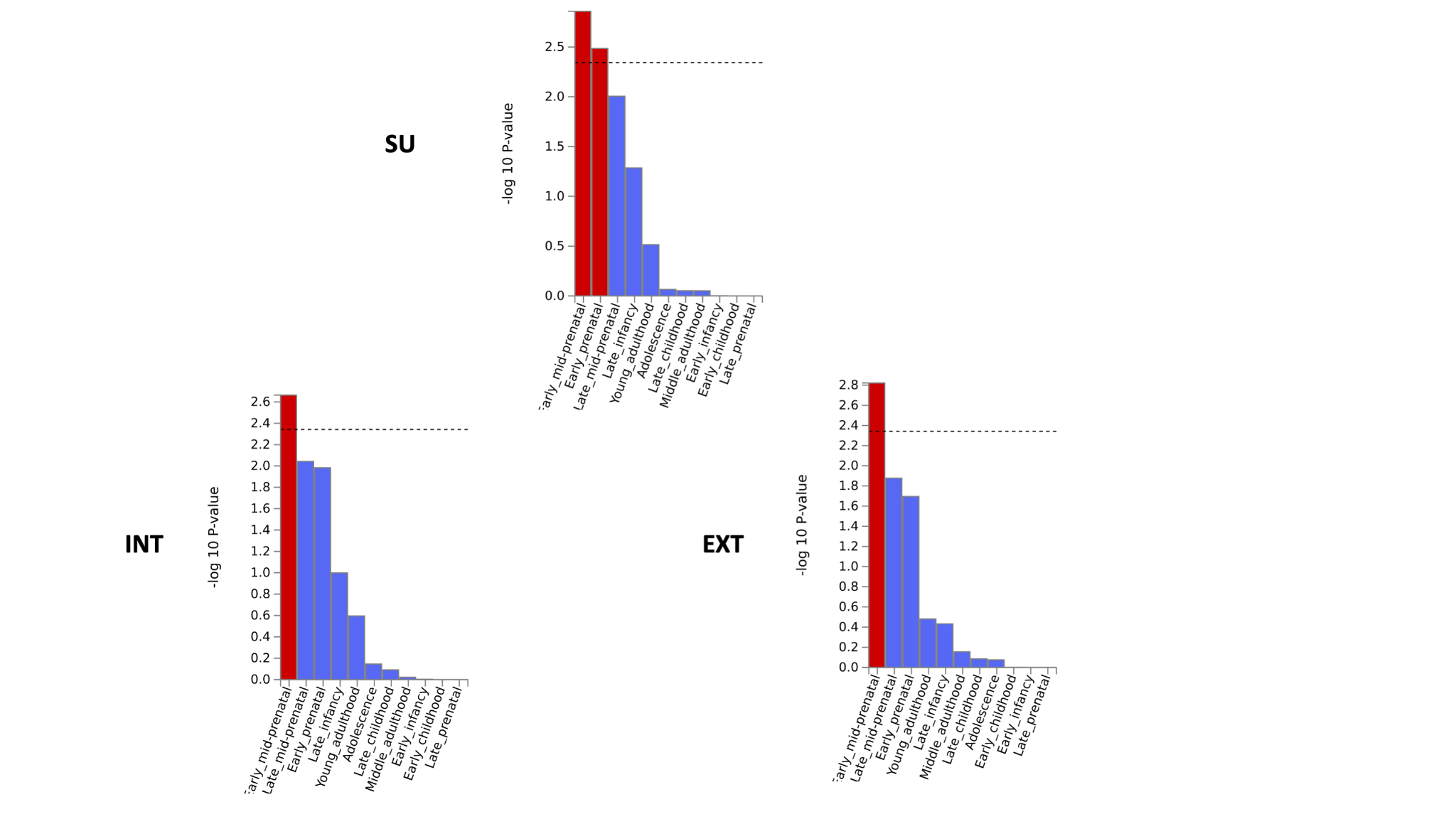

c. **Results from MAGMA tissue expression analysis using BrainSpan’s 11 general developmental stages of brain samples.**

**Supplementary Figure S3.** FUMA Results for three factors representing SU-related (SU), non-SU internalizing (INT), and non-SU externalizing genetic variance (EXT).

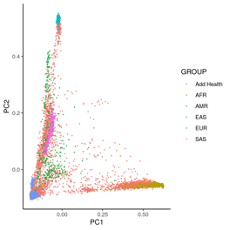

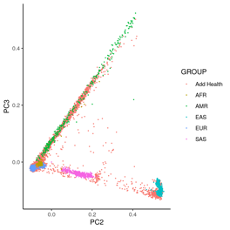

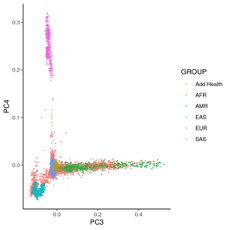

**Supplementary Figure S4.** Principal components (PCs) plots to determine ancestry using the 1000 Genome Reference Panel super populations. AFR = African; AMR = Americas; EAS = East Asian; EUR = European; SAS = South Asian.

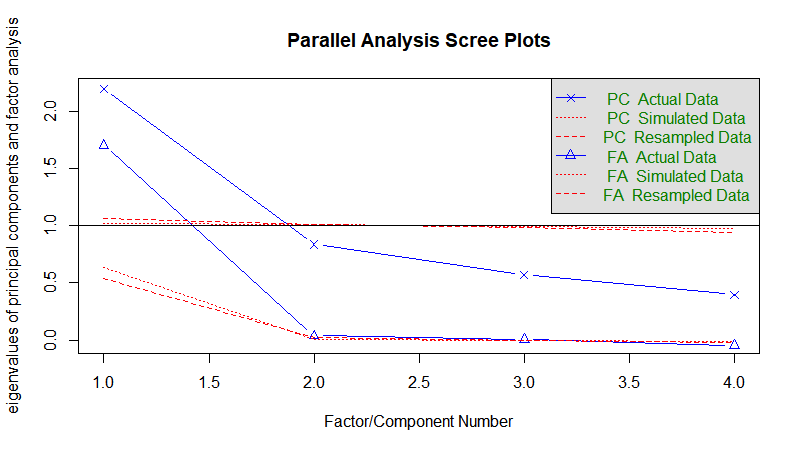

**Supplementary Figure S5.** Scree plot from parallel analysis of 30-day substance use variables supporting a one factor solution.
